# “General anxiety and depression are associated with the physical activity and social interaction levels: Study in Argentinean university students during the COVID-19 outbreak”

**DOI:** 10.1101/2022.01.07.22268803

**Authors:** Alejo Barbuzza, Pedro Benedetti, Celina Goyeneche, Victoria Reppucci, Franco Moscato, Daniela Ramirez Butavand, Cynthia Katche, Jorge Horacio Medina, Diego Moncada, Haydée Viola, Fabricio Ballarini

## Abstract

**Background:** Due to the coronavirus disease 2019 (COVID-19), the planet is going through a historical time of exceptional concern and uncertainty, which impacts people’s mental health. Here, we explored the levels of depression and Generalized Anxiety Disorder (GAD) and their relation with the degree of physical activity and social interaction during the pandemic.

**Methods:** We performed a structured survey containing the PHQ-9 and GAD-7 tests to evaluate depressive symptoms and GAD levels. We also asked about weekly physical activity and the level of social interaction. We surveyed two groups of University students in the Buenos Aires Metropolitan Area: an internal group from the *Instituto Tecnológico de Buenos Aires* (ITBA), and an external group of students from multiple universities. The survey was conducted in late October/early-November 2020, after a peak of contagions. Some of the participants were surveyed again in January 2021, during academic holidays and after a valley of contagion, for longitudinal analysis

**Results:** Our data show that men and women of both groups exhibited a significant positive linear correlation between depression and GAD levels. Moreover, low levels of depression and anxiety were associated with performing physical activity for more than two days a week and to longer periods of social interaction. Finally, the second survey revealed a decrease of the symptoms.

**Conclusions:** Our results suggest that performing regular physical activity and avoiding long periods of social isolation gave benefits to mental health. We suggest that public policies could consider protecting these behaviors under health and safety standards.

## Introduction

COVID 19 has a dramatic effect globally. Despite the primary clinical syndrome of COVID-19 being a respiratory disorder, it is already evident that the direct and indirect psychological and social effects of this pandemic are pervasive and could affect mental health now and in the future. The Generalized Anxiety Disorder (GAD) is one of the most common mental disorders. Depression is one of the most prevalent and treatable mental disorders and is regularly seen by a wide spectrum of healthcare providers, including mental health. There are many works that recorded the levels of GAD and depression during the COVID-19 pandemic. These levels were reported in studies attained to the general population from countries as diverse as China, Hong Kong, Greece, Argentina, Cypriot, Ireland, Austria, Germany and the US (Bäuerle et al., 2020; Choi et al., 2020; Fernández et al., 2020; Huang and Zhao, 2020; Hyland et al., 2020; Liu et al., 2020; Parlapani et al., 2020; Pieh et al., 2020; Solomou and Constantinidou, 2020; Wang et al., 2020). A major adverse consequence of the COVID19 pandemic is likely to be increased social isolation and loneliness which are strongly associated with anxiety and depression (Ben-Ezra et al., 2020; Brooks et al., 2020; Cao et al., 2020; Fernández et al., 2020; Marroquín et al., 2020; Rubin and Wessely, 2020; Tang et al., 2020; Tull et al., 2020.)

In our country, the government decreed a nationwide lockdown on 18th March 2020 (Boletín Oficial República Argentina, 2020a). These restrictive executive orders were renewed every two weeks according to the epidemiological situation until 7th November when the government decreed the end of the lockdown (Boletin Oficial República Argentina, 2020b). During this period only essential activities were permitted: essential shops were the only ones allowed to be open and individuals had permission to leave their homes exclusively for demonstrated necessities, such as health reasons, shopping for basic needs and essential work (Boletín Oficial República Argentina, 2020a). On 7th November 2020, the total confirmed cases of COVID-19 in Argentina were 1.236.851 with 33.348 confirmed deaths (Argentina.Gob.Ar, 2020a). From this point the restrictions began to decrease gradually and different activities were habilitated, for example team sports, bars, restaurants and tourism.

A recent publication made in our country highlighted the sudden environmental impacts on affective states a week after the start of the lockdown, even showing its greater impact on the young population (Torrente et al., 2021). Here, we are interested in evaluating the levels of GAD and depression at the time of closing the lockdown, after people have gone through almost 8 months of policies that strongly restricted social interaction. We are even interested in focusing the study on the population of university students since there are some inconsistent reports from different countries. In China, moderate-severe scores are only around 3% (Cao et al., 2020) and, at the other extreme, in Bangladesh, moderatesevere anxiety records show a prevalence greater than 80% (Dhar et al., 2020). Another study carried out in Germany between March and May 2020, yielded a percentage close to 25% of the population of young people between 18 and 24 years old (Bäuerle et al., 2020). Owing to the great variability of these percentages, we decided to carry out the work on university students in our country considering a sample from our university (Internal Group from ITBA) and another from various universities (External Group). Then, after a first survey at Oct-Nov 2020, some of the participants answered the survey again in January 2021, when the numbers of infected decreased. Thus, we were able to perform cross sectional and longitudinal analysis of mental health during specific pandemic periods.

As defined by Marroquin and colleagues (2020) the term “social distancing”, which could be considered opposite to social interaction, comprises interventions spanning both public and private levels. They included either “personal distancing” behaviors to reduce virus transmission (e.g., avoiding physical contact or close proximity with non-household members; reducing the use of shared public spaces) and government-imposed stay-at-home policies. Here, we asked the participants about the moment they started to interact with people who were not part of the home environment. That is, the moment of the year when they started to meet in social gatherings.

On the other hand, physical activity is defined as any bodily/corporal movement produced by skeletal muscles that results in energy expenditure (Caspersen et al., 1985). It was reported that walking, moderate, vigorous, and total physical activity levels have been reduced during the COVID-19 pandemic confinements in university students of different countries (López-Valenciano et al., 2021). Physical activity is known to aid psychological well-being, and it is a coping strategy during this pandemic. In a recent review, it was concluded that performing physical activity during COVID-19 is associated with less depression and anxiety (Wolf et al., 2021). In this work we survey in our university students the frequency that they perform physical activity and we contrasted it with the self-perceived levels of anxiety and depression.

## Material and Methods

### Participants, study design and procedure

Two different groups of university students between 18 and 30 years, residents of Metropolitan Area of Buenos Aires (AMBA), Argentina were recruited to participate in this study during the COVID-19 outbreak: 1_ an Internal Group was recruited within the Instituto Tecnológico de Buenos Aires, a Technological University; 2_ an External Group, more heterogeneous than the first one, recruited from students of several Universities and careers, through the social media. Using the Google Forms platform, both groups were surveyed with the GAD-7 and PHQ-9 tests, as well as a questionnaire regarding the number of days they performed physical activity and the period they started social interaction. Data was collected first in October/November 2020 (from 22nd October 2020 to 7th November 2020; Internal Group, 128 students, 51% women; external group 132 students 53% women) and it was repeated in January 2021 for both groups (from 6th January 2021 to 14th January 2021; Internal Group, 27 students, 62% women; External Group, 47 students, 61% women). The subgroup of students that answered in both periods was used for the longitudinal analysis. The AMBA is a geographical region composed by the Autonomous City of Buenos Aires and multiple political units of the Buenos Aires province, which together compose a continuous urban conglomerate of approximately 15 million people.

### Survey structure Measures

Participants responded about personal information in the first section of the survey. Both groups of university students informed their name, surname, area of residence (‘AMBA’ or ‘Not AMBA’), age (‘18’ up to ‘30’), gender (‘man’, ’woman’ and ‘other’), education level (‘undergraduate university’ or ‘graduated’) and other demographic characteristic. Then the survey inquired whether they formed physical activity, the grade of social interaction and about perceived levels of anxiety and depression.

### Physical Activity

All participants were asked if they performed physical activity during COVID-19 outbreak and how many times per week they normally did it (from ‘0’ to ‘7’).

### Social interaction information

Both groups were asked about the grade of social interaction, that is, when they started to see, in a non virtual way, people outside their cohabiting group, from ‘March 2020’ (month when restrictions began in Argentina) up to ‘November 2020’ in the first data collection and up to ‘January 2021’ in the second data collection. Data obtained was classified in three categories: Low social interaction (LSI, those people who did not practiced SI for 6 months or more) Medium social interaction (MSI, people who practiced SI between 3 to 5 months) and High social interaction (HSI, those who kept SI for 6 month or more).

### Generalized anxiety symptoms

Generalized anxiety was assessed through the 7-item Generalized Anxiety Disorder Scale (GAD7, Spitzer et al. 2006). This test measures symptoms over the last 2 weeks. It has been reported that GAD-7 showed strong reliability and validity in identifying probable DSM-IV generalized anxiety disorder (GAD; American Psychiatric Association, 1994), and also it was used in different populations (Löwe et al., 2008; Plummer et al., 2016). Briefly, respondents reported their symptoms using a 4-point Likert rating scale ranging from 0 (not at all) to 3 (almost every day) such that the total score ranges from 0 to 21. Scores of 0–4 are thought to represent minimal anxiety, 5–9 mild anxiety, 10–14 moderate anxiety, and 15–21 severe anxiety (Spitzer et al., 2006). The reliability in this study was: 1_ October/November 2020, α=0.89 for Internal Group and α=0.88 for External Group; 2_ January 2021, α=0.71 and α=0.88 for the Internal and External Groups, respectively.

### Depression symptoms

Depression was assessed through the Patient Health Questionnaire (PHQ-9; Kroenke et al., 2001). PHQ-9 is a reliable and widely validated measure for detecting depression symptoms (Beard et al., 2016; Kroenke et al., 2001; Titov et al., 2011). Its nine items are based on DSM-IV diagnostic criteria for major depressive disorder (American Psychiatric Association, 1994). Briefly, the participants reported their symptoms using a 4-point Likert rating scale ranging from 0 (not at all) to 3 (almost every day) such that the total score ranges from 0 to 27. Scores of 0–4 suggest minimal depression, 5–9 mild depression, 10–14 moderate depression, 15–19 moderately severe depression, and 20–27 severe depression (Kroenke et al. 2001). In this study reliability was: 1_ October/November 2020, α=0.85 for the Internal Group and α=0.74 for the External Group; 2_ January 2021, α=0.82 and α=0.85 for the Internal and External Groups, respectively).

### Ethical Considerations

This study was approved by the Students Department of the Instituto Tecnológico de Buenos Aires, with the consent of The University authorities who felt interested in the psychological impact of the COVID-19 pandemic in the students, in order to take actions directed to attenuate this impact. Before answering the survey, each participant was provided with an informed consent that had to be approved to participate in the study. All procedures performed in this study were in accordance with the ethical standards of the institutional and/or national research committee and with the Helsinki Declaration of 1975, as revised in 2008.

### Statistical analysis

All data was analyzed using GraphPad Prism® 8.0.1 software. First, we calculated descriptive statistics for the sample, expressed as counts and percentages (%) for non-continuous variables (men, women), and as means with standard error of the mean (SEM) for continuous variables (GAD-score from GAD-7, Depression score from PHQ-9 and mean day of weekly days of physical activity).

The correlation between GAD and depression during 2020 and 2021 was evaluated in the total population, and for male and female students, using a Pearson correlation analysis between these two mental health parameters. To assess the influence of gender and physical activity on states of GAD and Depression on both populations, a series of paired sample tests (2020 vs 2021) were conducted.

In all cases, the differences between physical activity days per week were evaluated conducting a one-tailed Mann-Whitney analysis. For all the analysis, the differences were considered significant when p < 0.05 (α=0.05).

## Results

Our results show that the self-perceived levels of depression registered in October-November 2020 (from now on 2020) positively correlated with those of GAD in both genders and in both groups of students (Fig 1 A-F). The percentage of people with either moderate or severe GAD was 39% and 48% and with moderate or severe depression 37% and 46% in the Internal and External groups of students, respectively.

**Figure 1.**
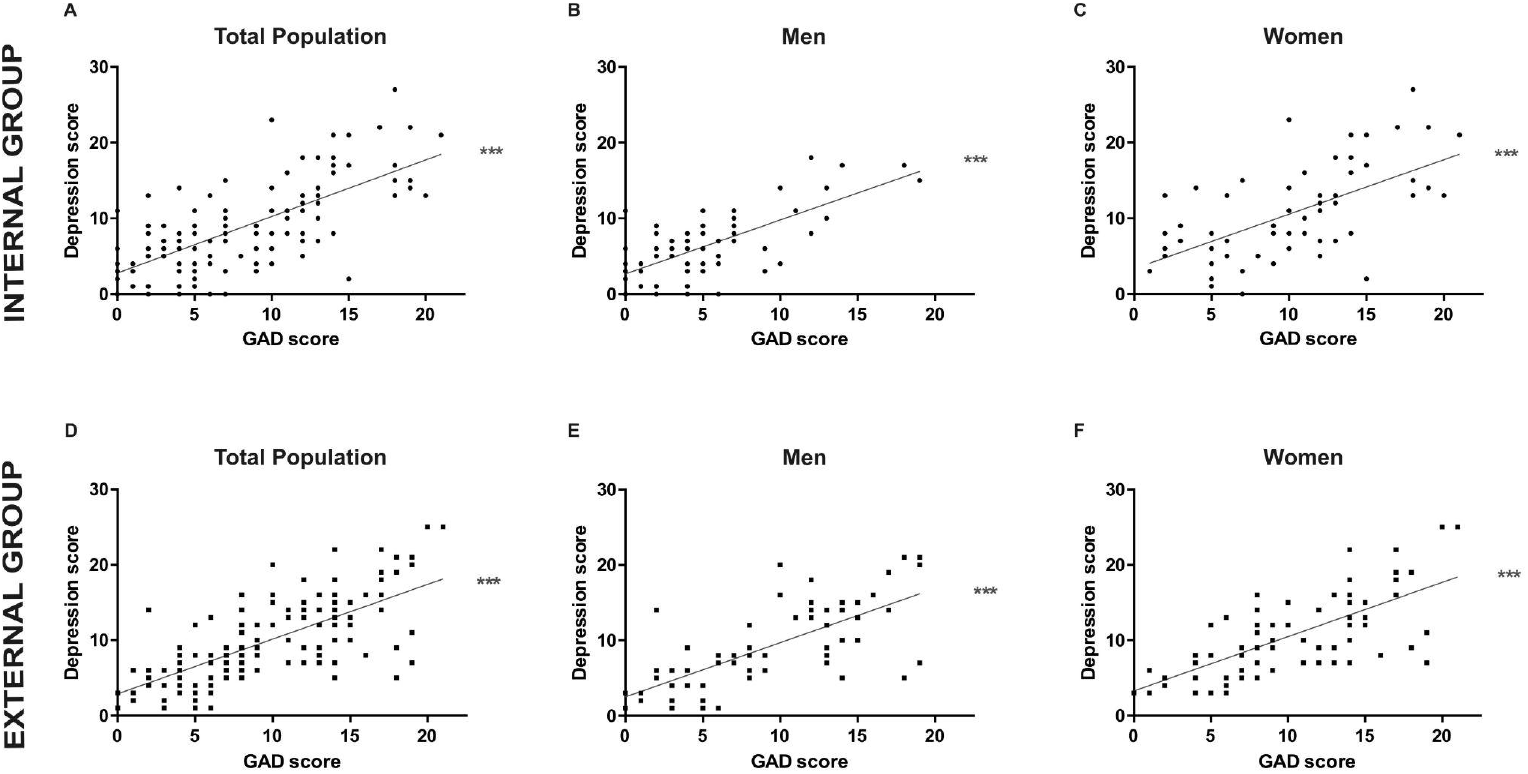
The levels of depression registered in 2020 positively correlate with those of GAD in both genders and populations of university students. ***Top panel:*** *Internal Group. Correlation between GAD and Depression scores in* ***(A)*** *the total population (n=128, r=0*.*703; *** p<0*.*001)*, ***(B)*** *the men population (n=63, r=0*.*704; *** p<0*.*001) and* ***(C)*** *the women population (n=65, r=0*.*619; p<0*.*001)*.). ***Bottom panel:*** *External Group. Correlation between GAD and Depression score in* ***(D)*** *the total population (n=132, r=0*.*703; ***p<0*.*001)*, ***(E)*** *the men population (n=61, r=0*.*709; *** p<0*.*001) and* ***(F)*** *the women population (n=71, r=0*.*696; *** p<0*.*001)*. In all cases a Pearson correlation analysis was performed.

By analyzing the relation between the frequency of physical activity and the self-perception of mental health, we observed that those individuals who performed exercise more than twice a week reported lower levels of GAD and depression, regardless they belonged to the Internal (Fig 2 A-B) or the External Group of students (Fig 2 E-F). In turn, we also observed a relation between the level of social interaction and mood. In this case, we observed that the Internal Group of students who kept high social interaction (HSI, those who sustained SI for six months or more) presented a significantly lower self-perception of GAD and depression than those who kept middle (MSI) and low (LSI) social interaction, (Fig 2 C-D). A similar, but not significant, trend to decrease was observed in the GAD levels of the students from External Group (Fig 2 G).

**Figure 2.**
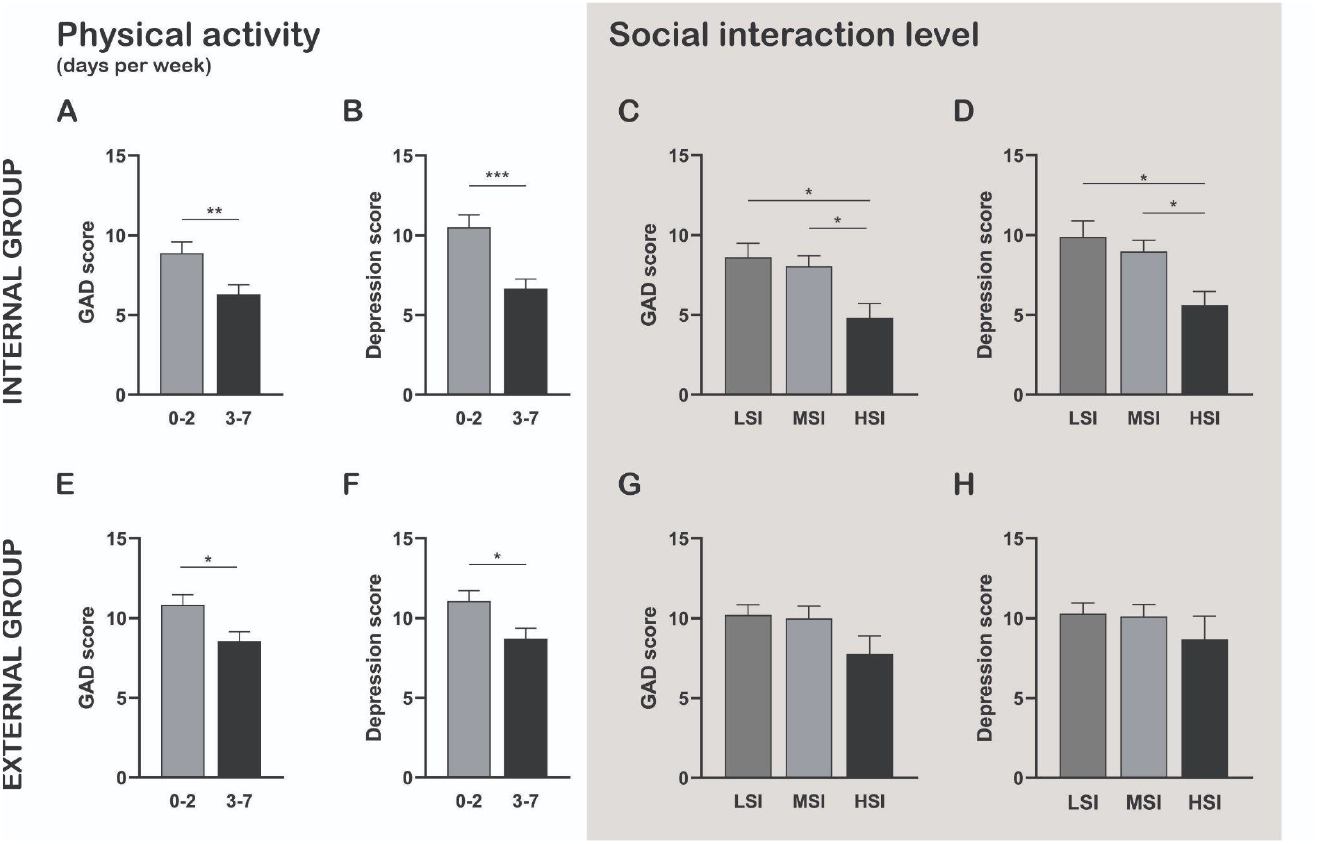
Higher physical activity frequency and longer periods of social interaction are associated with lower levels of GAD and depression. Figures show the level of GAD and Depression scores, depicted as mean +/- SEM, based on the days of physical activity per week (left panel) and the level of social interaction (right panel). **(A)** GAD and **(B)** Depression scores in the Internal Group of students who performed physical activity up to 2 days per week (gray bars, n=70-70, respectively) or between 3 to 7 (black bars, n=58); Student’s t-test, **p < 0.01, ***p < 0.001. **(C)** GAD and **(D)** Depression scores in the Internal Group of students who kept High (LSI, dark gray bars, n=47-47), Medium (MSI, light gray bars, n=59-59) or a HIgh (HSI, black bars, n=22-22) Social Interaction; Newman-Keuls analysis after One-way ANOVA, *p < 0.05. **(E)** GAD and **(F)** Depression scores in the External Group of students who performed physical activity up to 2 days per week (gray bars, n=74-74, respectively) or between 3 to 7 days (black bars, n=58); Student’s t-test, *p < 0.05. **(G)** GAD and **(H)** Depression scores in the External Group of students who kept Low (dark gray bars, n=67), Medium (light gray bars, n=49) or High (black bars, n=16) Social Interaction (p > 0.05).

A posterior gender analysis showed that GAD and depression levels in the men of the Internal Group of students was significantly lower than in the women (Fig 3 A-B). Applying this gender analysis to the level of physical activity and social interaction revealed that women of this group exercised less days than men (Fig 3 C; p<0.05), but kept equivalent levels of social interaction (Fig 3 D). Indeed, a contingency analysis revealed significant differences in the distribution of men and women according to the level of physical activity (p<0.05): High physical activity (60% men and 40% women); Low physical activity (39.66% men and 60,39% women). On the contrary, none of these differences were observed in the more heterogeneous External Group of students, where GAD and depression levels were equivalent between genders (Fig 3 E-F).

**Figure 3.**
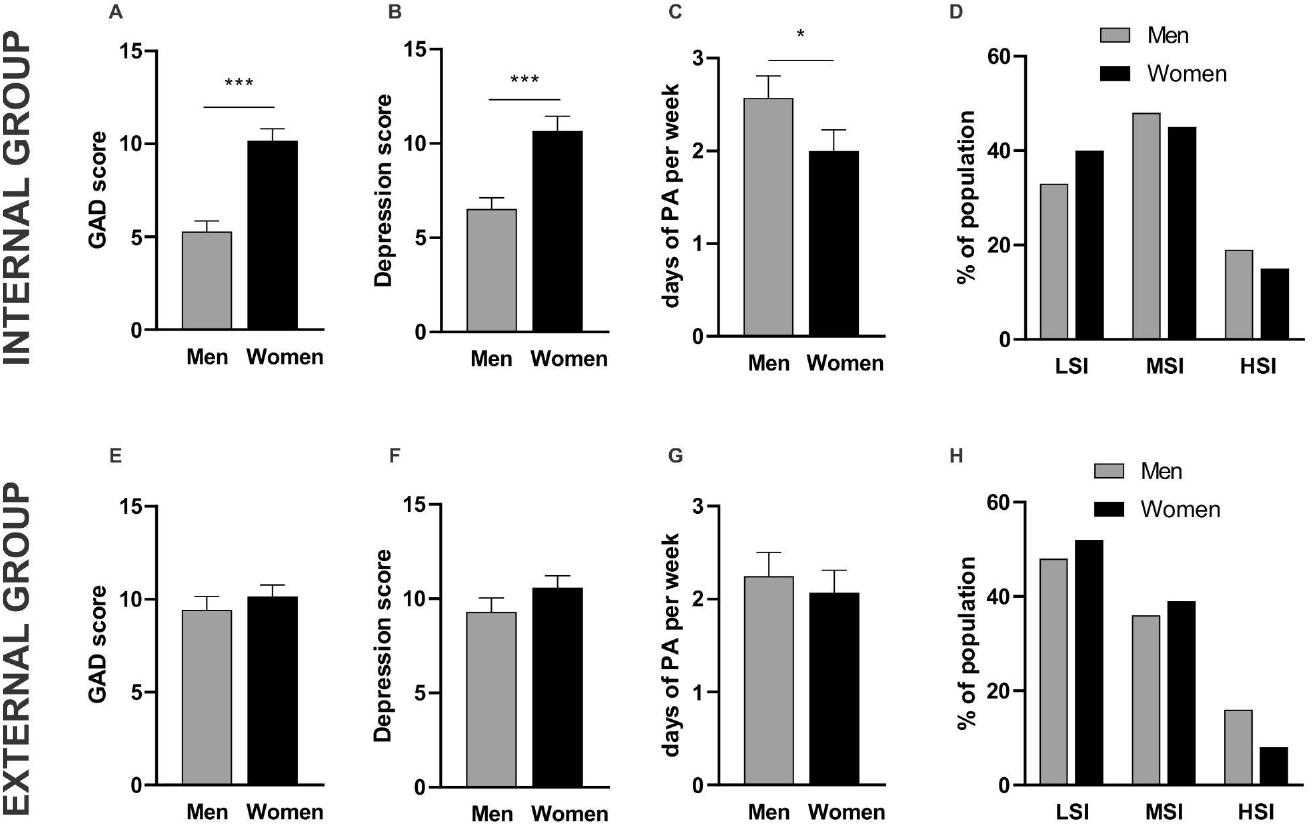
Men students of the Internal Group had lower levels of GAD and depression, while performing more days of physical activity, than women in the 2020. **Top panel:** Internal Group (men n=63, women n=65). Figures show **(A)** GAD levels (*** p<0.001, Student’s t-test), **(B)** Depression levels (*** p<0.001, Student’s t-test), and **(C)** day of physical activity (* p<0.05, one-tailed Mann-Whitney) expressed as mean +/- SEM. **(D)** the distribution of Social Interaction (as % of population. HLSI, MSI and HSI: Low, Medium and High Social Interaction, respectively. p mayor 0.05, Student’s t-test) between men (gray bars) and women (black bars). **Bottom panel:** External Group (men n=61, women n=71). Figures show **(E)** GAD levels (p>0.05 Student’s t-test), **(F)** Depression levels (p>0.05, Student’s t-test), and **(G)** day of physical activity (p>0.05, one-tailed Mann-Whitney) expressed as mean +/- SEM. **(H)** the distribution of Social Interaction (p>, 0.05 Student’s t-test) between men (gray bars) and women (black bars).

In January 2021 (from now on 2021) after a valley of infected people and in the valley of registered deaths, we repeated the survey. Here, the cross-sectional analysis confirmed the positive correlation between GAD and depression in both the Internal (Fig 4 A-C) and the External (Fig F-H) Groups of students. Nevertheless, this time the analysis by gender showed no differences between men and women in neither of the groups (Fig 4 D, E, I, J).

**Figure 4.**
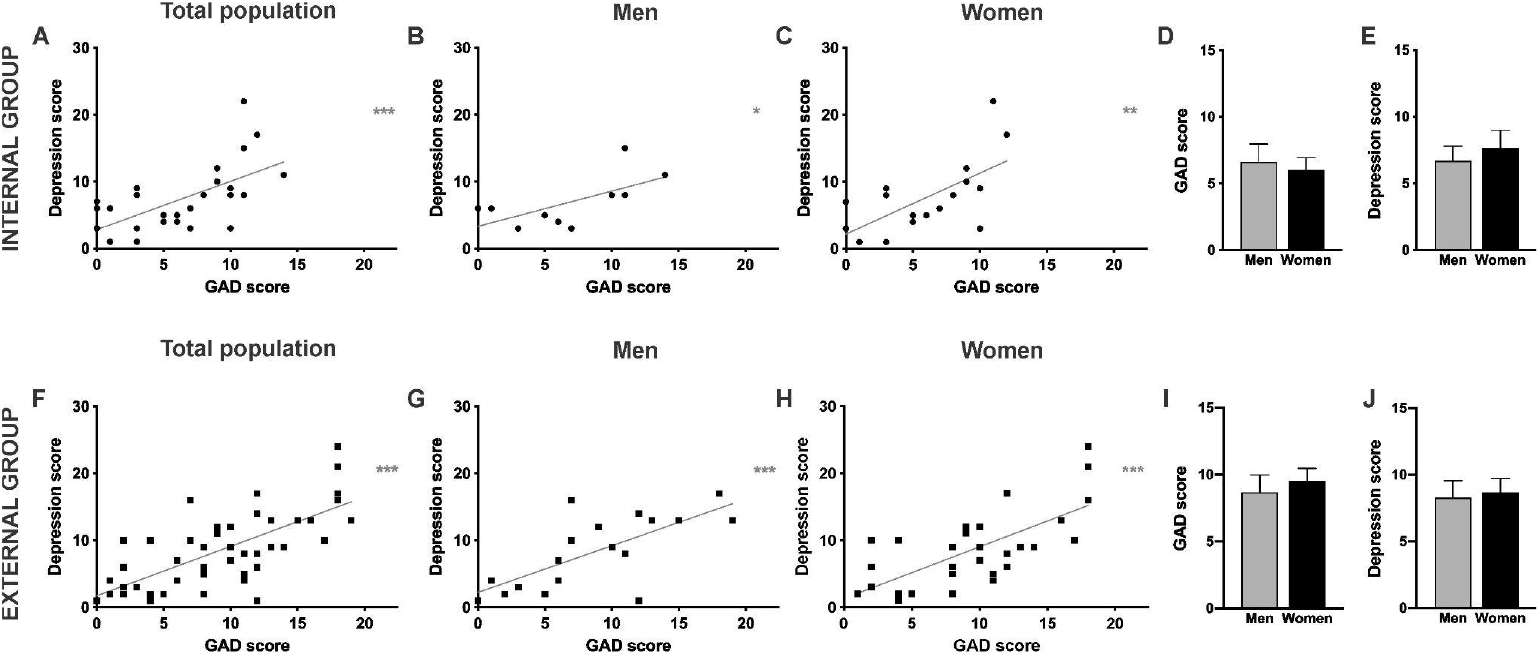
The levels of depression registered in 2021 also correlated positively with those of GAD in both genders and groups; besides the differences between genders in the Internal Group vanished. **Top panel:** Internal Group. Correlation between GAD and Depression scores in **(A)** total population (n=27, r=0,612, ***p < 0.001), **(B)** men population (n=10, r=0,647, *p < 0.01) and **(C)** women population (n=17, r=0,643, **p < 0.01). GAD **(D)** and Depression **(E)** scores of men (gray bars, n=10) and women (black bars, n=17) expressed as mean + SEM (Student’s t-test, p>0.05). **Bottom panel:** External Group. Correlation between GAD and Depression scores in **(F)** total population (n=47, r=0,701, ***p < 0.001), **(G)** men population (n=18, r=0,712, ***p < 0.001) and **(H)** women population (n=29, r=0,695, ***p <0.001). **(I)** GAD and **(J)** Depression scores of men (gray bars, n=18) and women (black bars, n=29) expressed as mean + SEM (Student’s t-test, p>0,05).

Finally, the longitudinal analysis, carried out with those students who answered the surveys twice, revealed that the levels of GAD and depression in 2021 were significantly lower than in 2020 for both the Internal Group (Fig. 5 A, D) and the External Group (Fig 6 A, D). In the particular case of the Internal Group, the decrease at the populational level was due to the lower records observed in women students (Fig 5 C, F), which also showed an increase in their weekly physical activity compared to the first survey (Fig 5 H). In contrast to women, the men of this group of students kept equivalent levels of GAD, depression and physical activity between the two periods (Fig 5 B, E, G). In the case of the External Group, the decrease in GAD and depression was observed in students of both genders (Fig 6 B, C, E, F). However, this decrease was not associated with changes in the frequency of physical activity (Fig 6 G, H).

**Figure 5.**
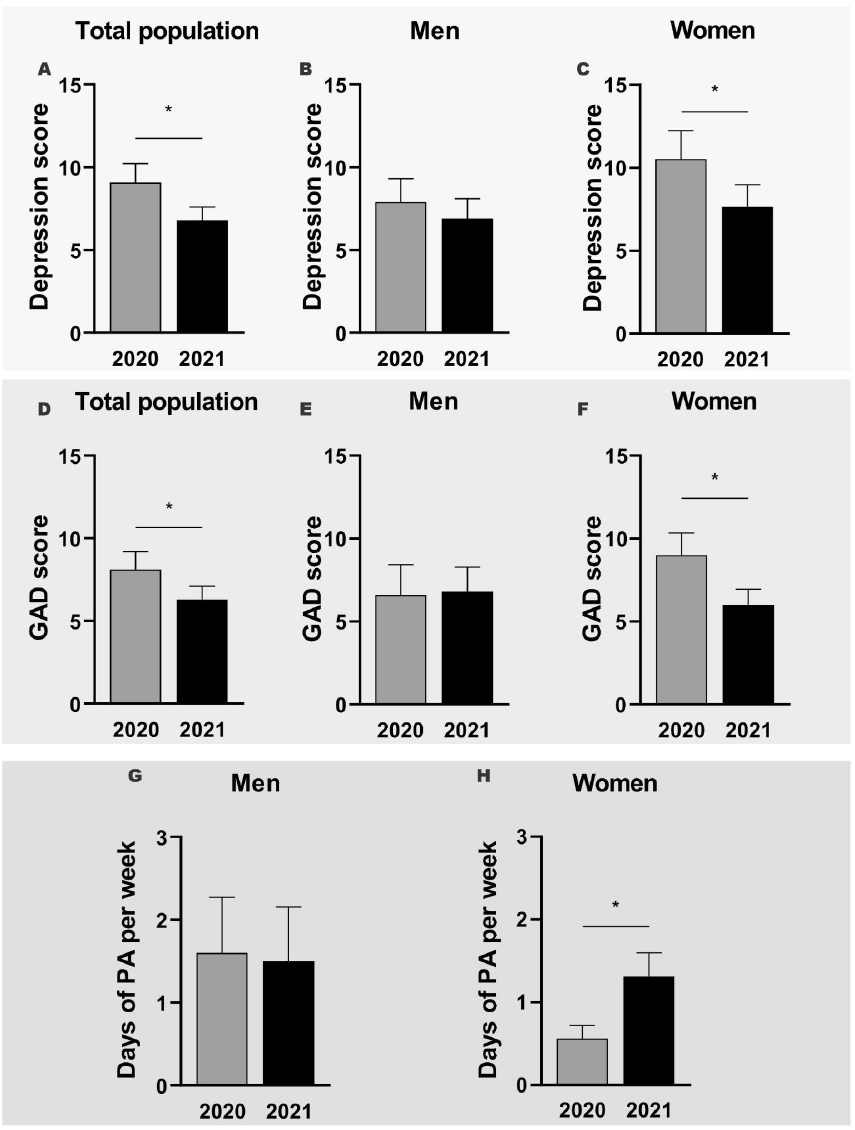
Longitudinal analysis of GAD score, depression level and days of physical activity in the Internal Group between 2020 and 2021. **Top Panel:** Depression scores for **(A)** the total population (n=26, an outlier was removed. Paired t-test, * p<0.05), **(B)** the men students (n=10. paired t-test, p>0.05) and **(C)** the women students (n=17, paired t-test, *p<0.05). **Medium Panel:** GAD score for **(D)** the total of students (n=27 paired t-test, p<0.05), **(E)** the men students (n=10, Paired t-test p>0.05) and **(F)** the women students (n=17, Paired t-test, *p<0.05). **Bottom panel:** days of physical activity per week in **(G)** the men (n= 10, paired t-test p>0.05) and **(H)** women (n=17, paired t-test ** p<0.01) students. All data is expressed as mean + SEM.

**Figure 6.**
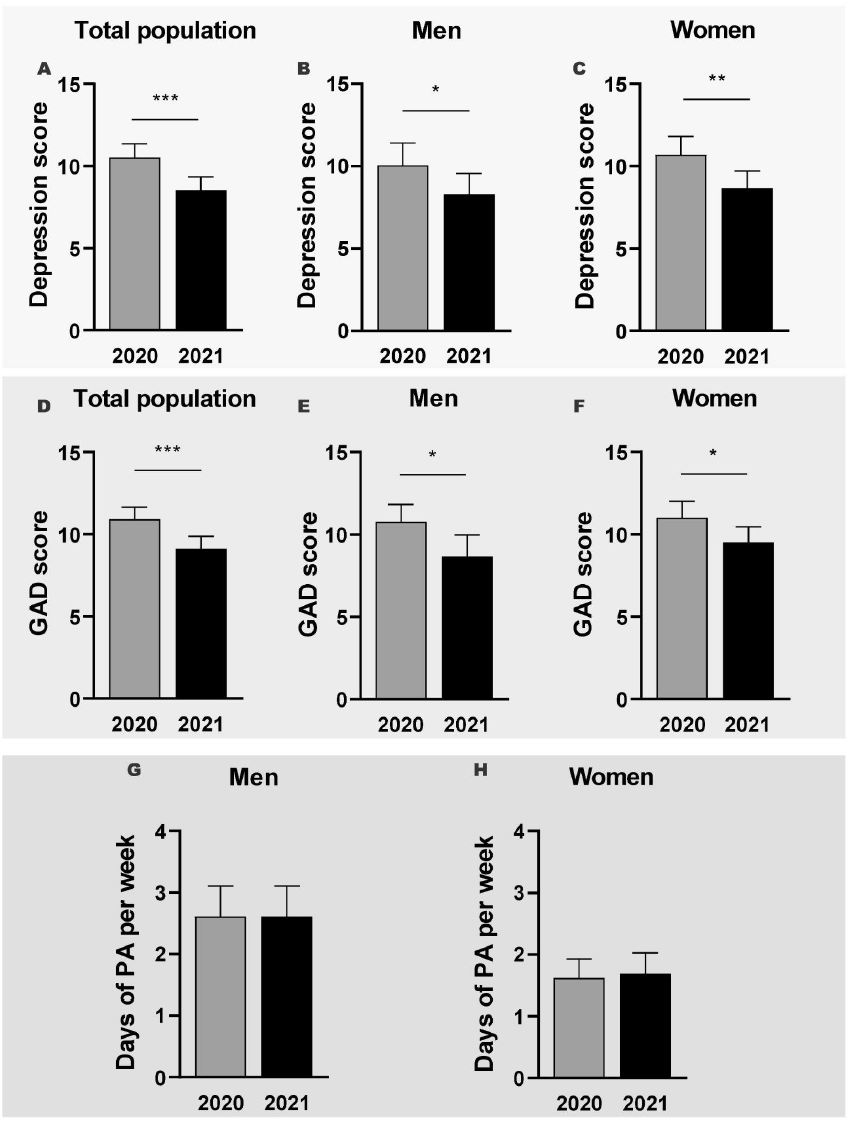
Longitudinal analysis of GAD scores, depression levels and days of physical activity in the External Group between 2020 and 2021. **Top Panel:** Depression scores for **(A)** the total population (n=47), Paired t-test, *** p<0.001) **(B)** the men students (n=18, Paired t-test, * p<0.05) and **(C)** the women students (n=29, Paired t-test, ** p<0.01). **Medium Panel:** GAD score for **(D)** the total of students (n=47, Paired t-test, *** p<0.001), **(E)** the men students (n=18, Paired t-test, * p<0.05) and **(F)** the women students (n=29, Paired t-test, * p<0.05. **Bottom panel:** days of physical activity per week in **(G)** the men (n=18) and **(H)** women students (n=29) (Paired t-test, p>0,05). Data are expressed as mean + SEM.

## Discussion

Depression and GAD represent distinct dimensions of the mental health state; however, it is frequent to see a high comorbidity in the symptoms of these disorders (Clark et al,. 1994). The main findings of the present study, performed in two groups of university students, revealed a significant positive linear regression between the levels of GAD and depression in both men and women. These data were obtained 8 months after the COVID-19 pandemic was declared, after a first peak of contagion and during the first peak of deaths in the Buenos Aires Metropolitan Area, Argentina. The percentage of the students with either moderate or severe GAD was 39% and 48% and with moderate or severe depression 37% and 46% in the Internal and External Groups, respectively. The percentage of subjects who declared social isolation of at least 6 months (low social interaction) was 37% in the Internal Group and 50% in the External one, while those who reported a frequency of physical activity greater than 2 times a week were 55% and 49%, respectively. Our analysis showed a significant decrease in the mean levels of GAD and depression associated with both: a physical activity frequency greater than twice a week as well as to high levels of social interaction. Finally, a longitudinal subsample of students, surveyed two months later when daily contagion cases had decreased by around 30% and the number of daily deaths was in a minimum, showed that their GAD and depression levels decreased significantly with respect to the first survey.

As a starting point, we want to contextualize our results considering the prevalence of moderate-severe levels of GAD and depression registered in our country from the beginning of the lockdown. In March 2020, a week after the lockdown began, a prevalence of 35% for GAD and 52% for depression was registered in a population of similar age to that of our study (Torrente et al., 2021). Then, a survey conducted in April 2020, showed that 31.8% of the subjects reported symptoms of anxiety and 27.5% of depression, although here it was recorded in adults between 18 to 92 years old (Fernandez et al., 2020). Also, there are data from April 2020 focusing on the study of depression and reported prevalence of 41% in young people aged 18 to 27 years, while in June 2020 it was reported 58% (Badellino et al., 2021). Here, the values registered for Oct-Nov 2020 in the university population was 39% and 48% for GAD and 37% and 46% for depression in the Internal and External Groups, respectively. These data as a whole suggest that in the young population the prevalence of GAD has shown a slight but sustained increase and that depression begins to show a slight decrease from the start of the lockdown.

In the present work, we also performed a longitudinal study in a subsample of participants. We observed that both depression and GAD mean scores significantly decreased in the second survey performed two months later, probably reflecting the decrease in the fatality rates and the holiday period that preceded that survey. On the other hand, a study conducted in the United States in participants surveyed to study these symptoms, earlier in the COVID-19 outbreak and a month later, showed an increase in GAD and depression levels. Personal distancing behavior was associated with these increases (Marroquin et al., 2020). Also, the prevalence of depression and GAD increases slightly in Spain, reaching 37%, 46% respectively, when the authors relieved these data first at the peak with approximately 8000 infections a day and then a month later with half of infections (Planchuelo-Gómez, 2020). Further studies are necessary to understand how the progress of the pandemic affects mental health.

There are multiple factors that could influence psychological distress (Fernandez et al., 2020; Stanton et al., 2020). In this work, we focused on two of these, the physical activity and the social interaction. Regular physical activity is a key health behavior related to a lower risk of mental disturbance (Raglin, 1990). Rebar et al. (2015), reported significant inverse associations between physical activity participation with depression and anxiety levels in their meta-analysis performed in 2015. A similar scenario is being observed during the COVID-19 pandemic. In that sense, Wolf et al. (2021) published a systematic review showing that those persons who reported a higher total time spent in moderate to vigorous physical activity also had 12% to 32% lower chances of presenting depressive symptoms and 15–34% of presenting anxiety. In addition, other studies carried out in adult people showed a significant association of negative changes in the activity scores with worse depression, anxiety and stress during 2020 (Meyer et al., 2020; Stanton et al., 2020).

Recent evidence shows that the amount of walking, the level of moderate or vigorous physical activity as well as total exercise performed, have been reduced during the COVID-19 pandemic confinements in 3.500 university students from eight different countries (LópezValenciano et al., 2021). In a study carried out in Italy, it was observed that in all age groups, there is a significant reduction of the total physical activity observed during COVID-19, even this change was more noticeable in men. Also, subjects classified as highly active and moderately active showed a significant correlation between the change in total activity and the Psychological General Well Being Index, which included life domains like anxiety and depression (Maugeri et al., 2020). Here, we observed in a group of Argentinean university students that performing physical activity more than twice a week was associated with a significant decrease in the average GAD and depression scores. Moreover, in the women of the Internal Group of Students, the longitudinal study showed that the decrease in the levels of these emotional states correlated with an increase in their physical activity levels. In line with our results, recent research performed a survey in Austria and showed that the groups with GAD and depression levels lower than 10 were enriched in subjects who practice physical activity more than twice a week, something that was not observed in the group with moderate-severe scores (Pieh et al., 2020). Even more, another study conducted in Spain revealed that people who performed daily exercise during two weeks before the surveys presented reduced levels of depression and stress, although this work was not focussed on university students (Planchuelo-Gómez et al.,2020). Our results show that a frequent physical activity is associated with lower scores in GAD and depression also in university students impacted by the pandemia.

Recently Polero et al. (2020) reviewed the recommendations of performing physical activity during the pandemic. According to these studies, and in agreement with the WHO recommendations, they suggested to perform at least 150 min of moderate-intensity or 75 min of vigorous physical activity per week. In particular, the neurobiological mechanisms by which physical activity affects the levels of anxiety, reactivity to stress, depression and mood, include the regulation of the hypothalamic-pituitary-adrenal axis, effects on the endogenous opioid system and the increase of the brain-derived neurotrophic factor level (Bodnar and Klein, 2005; Philips 2017; Rimmele et al., 2007). Thus, owing to the interruption of physical activity routines due to anti-contagion policies, it is worth noting the importance of promoting a change of behavior in the exercise habits, including online physical activity with friends or a virtual community, which might work in favor of social support to continue these activities.

The other factor considered in this work, which could influence psychological distress, was the social interaction. Our results suggested that the subjects who kept high non-virtual social interaction presented lower self-perception of GAD and depression than those of the groups with medium to low interaction levels. These changes were mild in the External Group of Students but marked in the Internal one (about 3 to 5 points in the scores). A survey performed by Marroquin et al. (2020) in 118 US participants showed increases in GAD and depression associated with personal distancing behavior. In that work, the absolute changes in the symptoms were statistically significant but relatively modest (about 1 point in the scores). Another work, showed that the risk of mental illness was higher among quarantined China people than among the not quarantined, even in unaffected areas (Tang et al., 2020). Elevated psychological distress was also found among quarantined Chinese respondents during March 2020, which represented the 12,5% of the sample (Ben-Ezra et al., 2020). Some of the difficulties that make the quarantine an anxious and depressive experience include the feeling of being confined, the separation from family, the disruption of regular social activities, the problem of not getting paid because of missed work, and uncertainty regarding contracting the disease (Blendon et al., 2004; Tang et al., 2020). So, after reviewing 24 studies, Brooks et al. (2020) warn about the potential psychological impact of the quarantine or the social isolation, alerting that their duration period should be limited to what is absolutely necessary in order to control the climax of the pandemic.

Finally, some evidence reveals a greater psychological impact of COVID-19 on women, showing higher levels of stress, anxiety, and depression, compared to men. The research of Fernandez et al. (2020) performed in Argentina supports this assertion. In our study, we observed differences between genders only in the Internal Group of students, where men declared less symptoms of GAD and depression than women in the survey from 2020. This result was associated with a higher proportion of men students performing high physical activity compared to women of the same group. On the other hand, a non-significative but notorious trend to maintain high social interaction was observed also in the men students of the Internal Group. In addition, it is possible that other factors unexplored during this study could also influence the difference in GAD and depression between genders. It is worth noting that the gender differences in GAD, depression, physical activity and social interaction were not observed in the External Group of students. A plausible explanation for this discrepancy may have originated from the differences in homogeneity between these groups. While the Internal Group was composed of confirmed students of a technical university located in Buenos Aires city, the External Group was composed of students of multiple universities, which could be public or private, that offer a wide range of careers (ie. from philosophy to medicine), and that are located in distinct sections of the Buenos Aires Metropolitan Area. Whereas studies conducted in Greek, Cypriot and Ireland communities found some mood sexual dimorphism (Hyland et al., 2020; Parlapani et al., 2020; Solomou and Constantinidou, 2020), other surveys performed in college students in China or in 1000 Australian people did not show such effects (Cao et al., 2020; Stanton et al., 2020). However, in those works a low average score for anxiety and depression was found.

In summary, our study shows that University students of the Buenos Aires Metropolitan Area in Argentina had values of GAD and depression that correlated in a linear and positive way in two different time periods of the COVID-19 pandemic. In addition, longitudinal analysis revealed that GAD and depression levels decreased in January 2021 when daily contagion cases had decreased and during a minimum of reported deaths. The analysis also revealed that a frequency of physical activity higher than twice a week is associated with a decrease in the average score of these emotional states. It also suggested that a higher social interaction is associated with decreased levels of GAD and depression. Based on our results, we argue that governments should consider the importance of physical exercise and the social interactions in mental health of university student, and therefore implement policies to promote these activities under health safety standards, such as social distancing in open spaces, to restrict the viral propagation.

## Data Availability

All data produced in the present study are available upon reasonable request to the authors

## References

Badellino, H., Gobbo, M.E., Torres, E., Aschieri, M.E., Biotti, M., Alvarez, V., Gigante, C., Cachiarelli, M., 2021. ‘ It ‘ s the economy, stupid ‘: Lessons of a longitudinal study of depression in Argentina. Int. J. Soc. Psychiatry. https://doi.org/10.1177/0020764021999687

Bäuerle, A., Teufel, M., Musche, V., Weismüller, B., Kohler, H., Hetkamp, M., Dörrie, N., Schweda, A., Skoda, E., 2020. Increased generalized anxiety, depression and distress during the COVID-19 pandemic: a cross-sectional study in Germany Alexander. J. Public Health (Bangkok). 1–7. https://doi.org/10.1093/pubmed/fdaa106

Beard, C., Hsu, K.J., Rifkin, L.S., Busch, A.B., Björgvinsson, T., 2016. Validation of the PHQ-9 in a psychiatric sample. J. Affect. Disord. 193, 267–273. https://doi.org/10.1016/j.jad.2015.12.075

Ben-Ezraa, M.,, Shaojing Sunb, Wai Kai Houc, R.G., 2020. The association of being in quarantine and related COVID-19 recommended and non-recommended behaviors with psychological distress in Chinese population. J. Affect. Disord. 275, 66–68. https://doi.org/10.1016/j.jad.2020.06.026

Blendon, R.J., Benson, J.M., Desroches, C.M., Raleigh, E., Taylor-clark, K., 2004. The Public ‘ s Response to Severe Acute Respiratory Syndrome in Toronto and the United States. Clin Infect Dis. 38, 925–931. https://doi.org/10.1086/382355

Bodnar, R.J., Klein, G.E., 2005. Endogenous opiates and behavior : 2004. Peptides 26, 2629–2711. https://doi.org/10.1016/j.peptides.2005.06.010

BOLETIN OFICIAL REPUBLICA ARGENTINA - AISLAMIENTO SOCIAL, PREVENTIVO Y OBLIGATORIO Y DISTANCIAMIENTO SOCIAL, PREVENTIVO Y OBLIGATORIO -Decreto 875/2020, 2020. https://www.boletinoficial.gob.ar/detalleAviso/primera/237062/20201107 (Accessed 7 November 2020).

BOLETIN OFICIAL REPUBLICA ARGENTINA - AISLAMIENTO SOCIAL PREVENTIVO Y OBLIGATORIO - Decreto 297/2020., 2020. http://www.boletinoficial.gob.ar/detalleAviso/primera/227042/20200320 (Accessed 18 March 2020).

Brooks, S.K., Webster, R.K., Smith, L.E., Woodland, L., Wessely, S., Greenberg, N., Rubin, G.J., 2020. Rapid Review The psychological impact of quarantine and how to reduce it : rapid review of the evidence. Lancet 6736. https://doi.org/10.1016/S0140-6736(20)30460-8

Caspersen, C.J., Powell, K. E., Christenson, G.M., 1985. Physical activity, exercise, and physical fitness: definitions and distinctions for health-related research. Public Heal. Rep 100, 126–131.

Cao, W., Fang, Z., Hou, G., Han, M., Xu, X., Dong, J., 2020. The psychological impact of the COVID-19 epidemic on college students in China. Psychiatry Res. 287, 112934. https://doi.org/10.1016/j.psychres.2020.112934

Chen, T.-C., Wang, C.-H., Huang, T.-H., Pan, C.-Y., Tsai, C.-L., Chen, F.-C., 2014. Impact of acute aerobic exercise and cardiorespiratory fitness on visuospatial attention performance and serum BDNF levels. Psychoneuroendocrinology 41, 121–131. https://doi.org/10.1016/j.psyneuen.2013.12.014

Choi, E.P.H.; Hui, B.P.H.; Wan, E.Y.., 2020. Depression and Anxiety in Hong Kong during COVID-19. Int. J. Environ. Res. Public Health 17, 37–40. https://doi.org/10.3390/ijerph17103740

Clark, D.A., Steer, R.A., Beck, A.T., 1994. Common and Specific Dimensions of Self-Reported Anxiety and Depression : Implications for the Cognitive and Tripartite Models. J. Abnorm. Psychol. 103, 645–654.

Dhar, B.K., Ayittey, F.K., Sarkar, S.M., 2020. Impact of COVID-19 on Psychology among the University Students. Glob. Challenges 4, 1–5. https://doi.org/10.1002/gch2.202000038

Fernández-prados, J.S., María, A., Martínez, M., Lozano-díaz, A., 2020. Impacts of COVID-19 Confinement among College Students : Life Satisfaction, Resilience and Social Capital Online. Int. J. Sociol. Educ. 79–104. https://doi.org/10.17583/rise.2020.5925

Huang, Y., Zhao, N., 2020. Generalized anxiety disorder, depressive symptoms and sleep quality during COVID-19 outbreak in China : a web-based cross-sectional survey. Psychiatry Res. 288, 112954. https://doi.org/10.1016/j.psychres.2020.112954

Hyland, P., Shevlin, M., McBride, O,. Murphy, J., Karatzias, T., Bentall, R P., Martinez, A., Vallières, F., 2020. Anxiety and depression in the Republic of Ireland during the COVID-19 pandemic. Acta Psychiatr. Scand. 142, 249–256. https://doi.org/10.1111/acps.13219

Kroenke, K., Spitzer, R.L., Williams, J.B.W., 2001. The PHQ-9. J Gen Intern Med. 16, 606–613. https://doi.org/10.1046/j.1525-1497.2001.016009606.x

Liu, C.H., Zhang, E., Tin, G., Ba, W., Hyun, S., Chris, H., 2020. Factors associated with depression, anxiety, and PTSD symptomatology during the COVID-19 pandemic : Clinical implications for U. S. young adult mental health. Psychiatry Res. 290. https://doi.org/10.1016/j.psychres.2020.113172

López-valenciano, A., Suárez-iglesias, D., Sanchez-lastra, M.A., 2021. Impact of COVID-19 Pandemic on University Students ‘ Physical Activity Levels : An Early Systematic Review. Front. Psychol. 11, 1–10. https://doi.org/10.3389/fpsyg.2020.624567

Löwe B, Decker O, Müller S, Brähler E, Schellberg D, Herzog W, H.P., 2008. Validation and Standardization of the Generalized Anxiety Disorder Screener (GAD-7) in the General Population. Med Care 46, 266–274. https://doi.org/10.1097/mlr.0b013e318160d093

Marroquín, B., Vine, V., Morgan, R., 2020. Mental health during the COVID-19 pandemic : Effects of stay-at-home policies, social distancing behavior, and social resources. Psychiatry Res. 293, 113419. https://doi.org/10.1016/j.psychres.2020.113419

Maugeri, G., Castrogiovanni, P., Battaglia, G., Pippi, R., Agata, V.D., Palma, A., Di, M., Musumeci, G., 2020. Heliyon The impact of physical activity on psychological health during Covid-19 pandemic in Italy. Heliyon 6, e04315. https://doi.org/10.1016/j.heliyon.2020.e04315

Meyer, J., Mcdowell, C., Lansing, J., Brower, C., Smith, L., Tully, M., Herring, M., 2020. Changes in Physical Activity and Sedentary Behavior in Response to COVID-19 and Their Associations with Mental Health in 3052 US Adults. Int. J. Environ. Res. Public Health 17(18), 64. https://doi.org/10.3390/ijerph17186469

Ministerio de Salud, A., 2020. Noviembre de 2020. (2020a, November 1). Argentina.Gob.Ar, 2020. https://www.argentina.gob.ar/coronavirus/informes-diarios/reportes/noviembre2020%0A (Accessed 7 November 2020).

Parlapani, E., Holeva, V., Voitsidis, P., Blekas, A., Gliatas, I., Porfyri, G.N., Golemis, A., Papadopoulou, K., Dimitriadou, A., Chatzigeorgiou, A.F., Bairachtari, V., Skoupra, M., 2020. Psychological and Behavioral Responses to the COVID-19 Pandemic in Greece. Front. Psychol. 11, 1–17. https://doi.org/10.3389/fpsyt.2020.00821

Phillips, C., 2017. Review Article Brain-Derived Neurotrophic Factor, Depression, and Physical Activity : Making the Neuroplastic Connection. Neural Plast. 2017. https://doi.org/10.1155/2017/7260130Review

Pieh, C., Budimir, S., Probst, T., 2020. The effect of age, gender, income, work, and physical activity on mental health during coronavirus disease (COVID-19) lockdown in Austria. J. Psychosom. Res. 136, 110186. https://doi.org/10.1016/j.jpsychores.2020.110186

Planchuelo-gómez, Á., Odriozola-gonzález, P., Jesús, M., Luis-garcía, R. De, 2020. Longitudinal evaluation of the psychological impact of the COVID-19 crisis in Spain. J. Affect. Disord. 277, 842–849. https://doi.org/10.1016/j.jad.2020.09.018

Plummer, F., Sc, M., Manea, L., Sc, M., Psych, M.R.C., Trepel, D., Ph, D., Mcmillan, D., Ph, D., Psy, D.C., 2016. Screening for anxiety disorders with the GAD-7 and GAD-2 : a systematic review and diagnostic metaanalysis ☆. Gen. Hosp. Psychiatry 39, 24–31. https://doi.org/10.1016/j.genhosppsych.2015.11.005

Polero, P., Rebollo-seco, C., Adsuar, J.C., Jorge, P., Rojo-ramos, J., Manzano-redondo, F., Garcia-gordillo, M.Á., Carlos-vivas, J., 2021. Physical Activity Recommendations during COVID-19 : Narrative Review. Int. J. Environ. Res. Public Health 18, 65. https://doi.org/10.3390/ijerph18010065

Raglin, J.S., 1990. Exercise and Mental Health Beneficial and Detrimental Effects. Sport. Med. 9, 323–329. https://doi.org/10.2165/00007256-199009060-00001

Rebar, A.L., Stanton, R., Geard, D., Short, C., Duncan, M.J., 2015. A Meta-Meta-Analysis of the effect of physical activity on depression and anxiety in non-clinical adult populations. Health Psychol. Rev. 37–41. https://doi.org/10.1080/17437199.2015.1022901

Rimmele, U., Costa, B., Marti, B., Seiler, R., Mohiyeddini, C., Ehlert, U., Heinrichs, M., 2007. Trained men show lower cortisol, heart rate and psychological responses to psychosocial stress compared with untrained men. Psychoneuroendocrinology 627–635. https://doi.org/10.1016/j.psyneuen.2007.04.005

Rubin, G.J., Wessely, S., 2020. The psychological effects of quarantining a city. BMJ 313, 1–2. https://doi.org/10.1136/bmj.m313

Solomou, I., Constantinidou, F., 2020. Prevalence and Predictors of Anxiety and Depression Symptoms during the COVID-19 Pandemic and Compliance with Precautionary Measures : Age and Sex Matter. Int. J. Environ. Res. Public Health 2, 1–19. https://doi.org/doi:10.3390/ijerph17144924

Spitzer, R.L., Kroenke, K., Williams, J.B.W., Lo, B., 2015. A Brief Measure for Assessing Generalized Anxiety Disorder. Arch Intern Med 166, 1092–1097. https://doi.org/10.1001/archinte.166.10.1092

Stanton R, To QG, Khalesi S, Williams SL, Alley SJ, Thwaite TL, Fenning AS V.C., 2020. Depression, Anxiety and Stress during COVID-19 : Associations with Changes in Physical Activity, Sleep, Tobacco and Alcohol Use in Australian Adults. Int. J. Environ. Res. Public Health 17, 1–13. https://doi.org/10.3390/ijerph17114065

Tang, F., Liang, J., Zhang, H., Mohammedosman, M., He, Q., Wang, P., 2020. COVID-19 related depression and anxiety among quarantined respondents. Psychol. Health 0, 1–15. https://doi.org/10.1080/08870446.2020.1782410

Titov, N., Mcmillan, D., 2011. Psychometric Comparison of the PHQ-9 and BDI-II for Measuring Response during Treatment of Depression. Cogn. Behav. Ther. 40, 126–136. https://doi.org/10.1080/16506073.2010.550059

Torrente, F., Yoris, A., Low, D.M., Lopez, P., Bekinschtein, P., Manes, F., 2021. Sooner than you think : A very early affective reaction to the COVID-19 pandemic and quarantine in Argentina. J. Affect. Disord. 282, 495–503. https://doi.org/10.1016/j.jad.2020.12.124

Wang, C., Pan, R., Wan, X., Tan, Y., Xu, L., Ho, C.S., Ho, R.C., 2019. Immediate Psychological Responses and Associated Factors during the Initial Stage of the 2019 Coronavirus Disease (COVID-19) Epidemic among the General Population in China. Int. J. Environ. Res. Public Health 17, 17–29. https://doi.org/10.3390/ijerph17051729

Wolf, S., Seiffer, B., Zeibig, J.M., Welkerling, J., Brokmeier, L., Atrott, B., Ehring, T., Barreto, F., 2021. Is Physical Activity Associated with Less Depression and Anxiety During the COVID - 19 Pandemic ? A Rapid Systematic Review. Sport. Med. April 22, 1–13. https://doi.org/10.1007/s40279-021-01468-z

